# Development and use of a method based on the anti-N reactivity of longitudinal samples to better estimate SARS-CoV-2 seroprevalence in a vaccinated population

**DOI:** 10.1101/2022.08.15.22278798

**Authors:** Renée Bazin, Samuel Rochette, Josée Perreault, Marie-Josée Fournier, Yves Grégoire, Amélie Boivin, Antoine Lewin, Marc Germain, Christian Renaud

## Abstract

**Background:** Emerging evidence suggests that COVID-19 vaccination decreases the sensitivity of anti-nucleocapsid (N) serologies, making them less reliable to assess recently-acquired infections. We therefore developed and tested a new approach based on the ratio of the anti-N absorbance of longitudinal samples to overcome this limitation.

**Methods:** Previously vaccinated repeat plasma donors provided at least one pre-infection (reference) and one post-infection (test) sample. All samples were tested using an in-house anti-N ELISA. Seropositivity was determined based on the ratio between the anti-N absorbance of the test and reference samples. The ratio approach was tested in a real-world setting during three cross-sectional serosurveys carried out among plasma donors in Québec, Canada.

**Results:** Using a cut-off ratio of 1.5, the approach had a sensitivity of 95.2% among the 248 previously vaccinated and infected donors compared with 63.3% for the conventional approach. When tested in a real-world setting, the ratio-based approach yielded an adjusted seroprevalence of 27.4% (95% confidence interval [CI]=23.8%-30.9%) at the latest time point considered, compared to 15.1% (95% CI=12.2%-18.0%) for the conventional approach.

**Conclusions:** This article describes a new and highly-sensitive approach that captures a significantly greater proportion of vaccinated individuals with a recent history of SARS-CoV-2 infection.

## BACKGROUND

Since the beginning of the SARS-CoV-2 pandemic, serosurveys have been key to public health decision-making, but their conduct and interpretation have become more challenging. Seroprevalence may no longer be viewed as an indicator of progression to a putative state of herd immunity — a prospect that looks increasingly dim with the continual emergence of immune-escape variants.[1,2] Early serosurveys assessed anti-spike (S) or anti-receptor binding domain (RBD) seropositivity in vaccine-naïve individuals, but this approach cannot disentangle infection-from vaccine-induced seropositivity in vaccinated individuals. Anti-nucleocapsid (N) seroprevalence was initially viewed as a solution to this challenge but is more affected by seroreversion than anti-S or anti-RBD seroprevalence,[3–12] thus leading to declining or stagnating seroprevalence estimates despite rising case counts.[3,13,14] What is more, emerging evidence suggests that vaccination hinders the sensitivity of conventional anti-N assays. In a study of 4000 fully vaccinated health care workers, 23 experienced a breakthrough infection, but only six (26%) were seropositive for anti-N.[15] More recently, a study of COVE (i.e., mRNA-1273 vs. placebo to prevent COVID-19) participants with a confirmed history of SARS-CoV-2 breakthrough infection found that only 40.4% of vaccine recipients were seropositive for anti-N, as compared with 93.4% among placebo recipients.[16] Although preliminary, this evidence is worrisome as it may invalidate conventional anti-N assays as a marker of SARS-CoV-2 infections, leaving few alternatives to assess the progression of the pandemic.

The reduced rate of anti-N seroconversion among vaccinated individuals might be explained by reduced exposure to the N antigen following infection,[16] which may lead to anti-N levels that do not pass the seropositivity threshold of conventional anti-N assays. We postulated that this limitation could be addressed by comparing the anti-N levels of samples collected at different time points, thus enabling the detection of meaningful signal increases in response to recently acquired infections. Based on this premise, we sought to develop and test an empirical approach to overcome the issue of vaccination in anti-N serosurveys.

## METHODS

### Donors

The approach was developed and tested based on longitudinally collected plasma samples from regular plasma donors who consented to participate in a COVID-19-dedicated biobank in Québec, Canada (“PlasCov”; see **Supplementary Methods** for details on the biobank). The PlasCov biobank and the use of samples for the present study were approved by the Héma-Québec Research Ethics Board. To develop the approach, plasma samples from donors who met the following criteria were selected: (1) received ≥1 vaccine dose against SARS-CoV-2; (2) had a PCR-confirmed SARS-CoV-2 infection between 12/15/2021 and 03/20/2022 (i.e., during the Omicron wave); and (3) made ≥2 donations: one before — but as close as possible — to 12/15/2021 (i.e., the reference sample collected before the Omicron wave) and one between 01/02/2022 and 04/20/2022 after the confirmed infection (i.e., the test sample collected during the Omicron wave; see **Supplementary Methods** for other inclusion criteria). To test the approach in a real-world setting, we conducted three cross-sectional serosurveys in Québec (Canada) that included plasma donors who made ≥1 donation in one of the following periods (all during the Omicron wave): 01/17/2022-01/18/2022, 02/14/2022-02/15/2022, and 03/16/2022-03/18/2022 (see **Supplementary Methods** for other inclusion criteria). All of these donors were included in the analysis conducted with the conventional approach, and only those with ≥1 reference sample and ≥1 test sample were included in the analysis conducted with the new ratio-based approach (described further down).

### Anti-N ELISA

All samples used to develop and test the ratio-based approach were analysed for anti-N seropositivity using an in-house developed conventional anti-N ELISA. The assay was similar to a previously described anti-RBD assay,[17] except that the recombinant N antigen (Centre National en Électrochimie et en Technologies Environementales Inc., Shawinigan, Canada) was used (0.25 µg/ml) in lieu of the RBD antigen (2.5 µg/ml). This assay has a sensitivity of 98.5% and a specificity of 98.1% among unvaccinated and recently infected individuals using an absorbance cut-off value of 0.350 (see **Supplementary Methods** for details).

### Ratio-based approach to determine anti-N seropositivity

The herein described ratio-based approach determines anti-N seropositivity based on the ratio between the absorbances of the test and reference samples (anti-N ratio). The positivity cut-off for the anti-N ratio was selected based on the proportion of vaccinated donors with a prior confirmed infection being captured (i.e., sensitivity). A baseline threshold was applied to exclude samples with low absorbance values (i.e., falling in the assay’s background noise) and hence reduce false positives resulting from the technical variability of the assay. Donors for whom the test sample had an absorbance value below this threshold were considered seronegative and no ratio was calculated.

### Statistical analysis

Seroprevalence estimates were reported along with binomial proportion confidence intervals (CI) after adjusting for age, sex, and regional distribution (based on 2011 census data) for both cumulative and ratio-based approaches.

## RESULTS

### Development of the ratio-based approach

The approach was developed using 248 vaccinated donors with a PCR-confirmed infection during the Omicron wave (mean age: 40.6 years; females: 48.3%), including 246 (99.2%) who had received ≥2 vaccine doses before their infection. The median interval between the PCR result and the collection of the test sample was 31 days (interquartile range: 19-46 days).

The target sensitivity of the ratio-based approach was 95%, as recommended by national regulatory authorities.[18] A sensitivity > 95% was achieved with anti-N ratios of up to 1.5 and dropped abruptly at higher ratios (**Figure 1**). Therefore, a positivity cut-off of 1.5 was selected.

**Figure 1.**
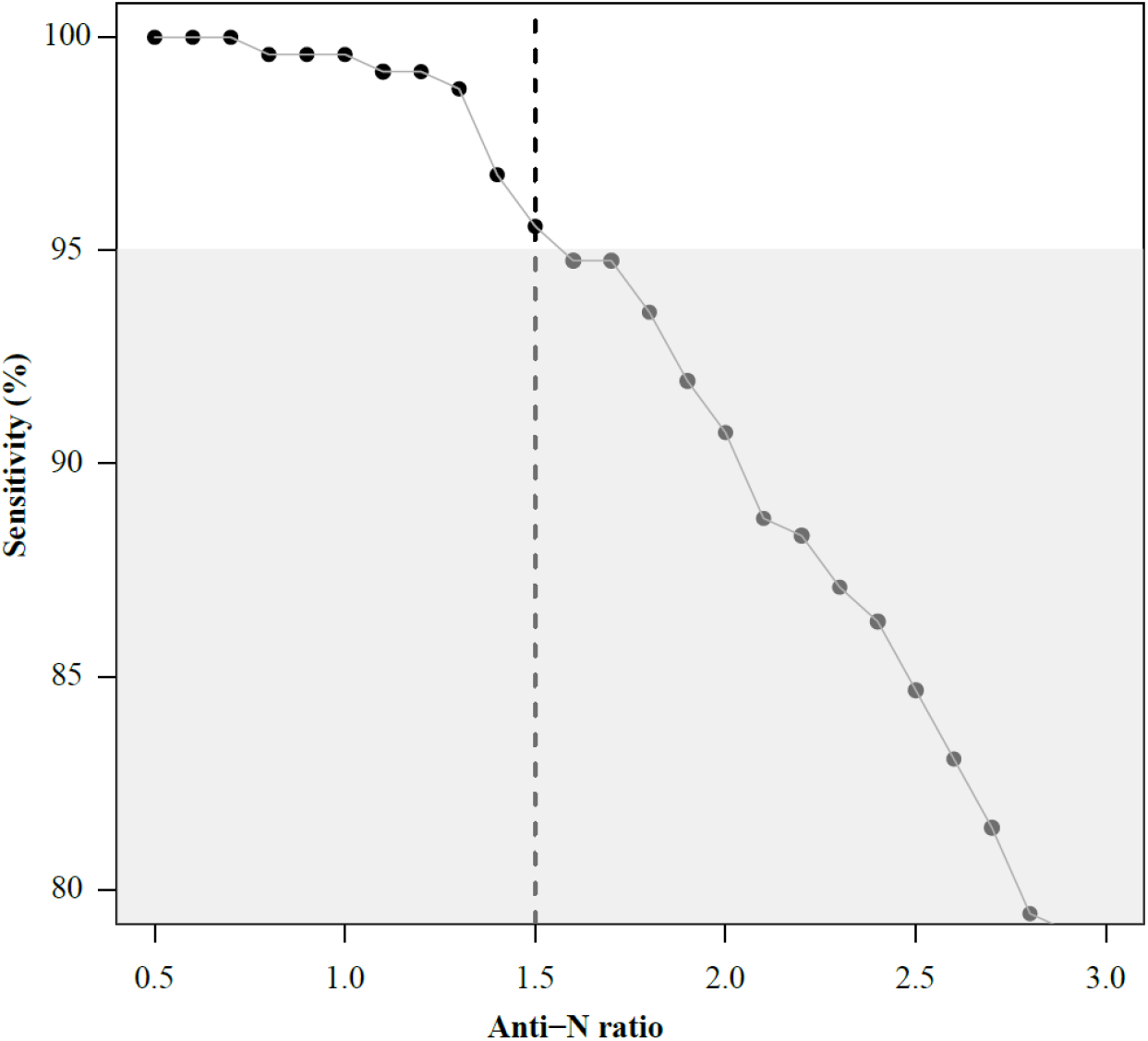
Sensitivity of detection of seropositive individuals captured using various anti-N ratio. The ratio of the anti-N signal between the test and reference samples was calculated. The sensitivity of detection obtained for ratios between 0.5 and 2.8 is shown. Sensitivity below the target value of 95% is indicated by the grey area.

To improve the specificity of the approach, we determined a limit of absorbance below which a test sample would not be included since low absorbance values are more variable and may inadvertently produce anti-N ratios ≥1.5 owing to the technical variability of the assay. Indeed, the coefficient of variation of 38 technical replicates of a negative control sample (pre-pandemic plasma pool) was 12.4% with a mean absorbance of 0.121. We therefore conservatively selected 0.100 for this lower limit of absorbance in test samples.

Applying the above-mentioned thresholds, the sensitivity of the ratio-based approach to identify new infections was estimated at 95.2% (236/248), whereas that of the conventional approach (based on the 0.350 seropositivity cut-off of the anti-N ELISA at the post-infection time point) was 63.3% (157/248).

The stability of the baseline anti-N signal in reference samples was assessed among seven donors with an anti-N ratio ≥1.5 and three reference samples. For these donors, the anti-N signal of the reference samples had a mean coefficient of variation (CV) of 10.7% (**Figure 2**), and all had an anti-N ratio ≥1.5 regardless of the reference sample considered. The analysis of an additional 12 donors with 2 reference samples confirmed the low variability of the baseline anti-N signal, with a mean CV of 8.4% (data not shown). False positives due to an unstable anti-N signal in reference samples thus appear to be infrequent, although a larger sample would be needed to better estimate this rate.

**Figure 2:**
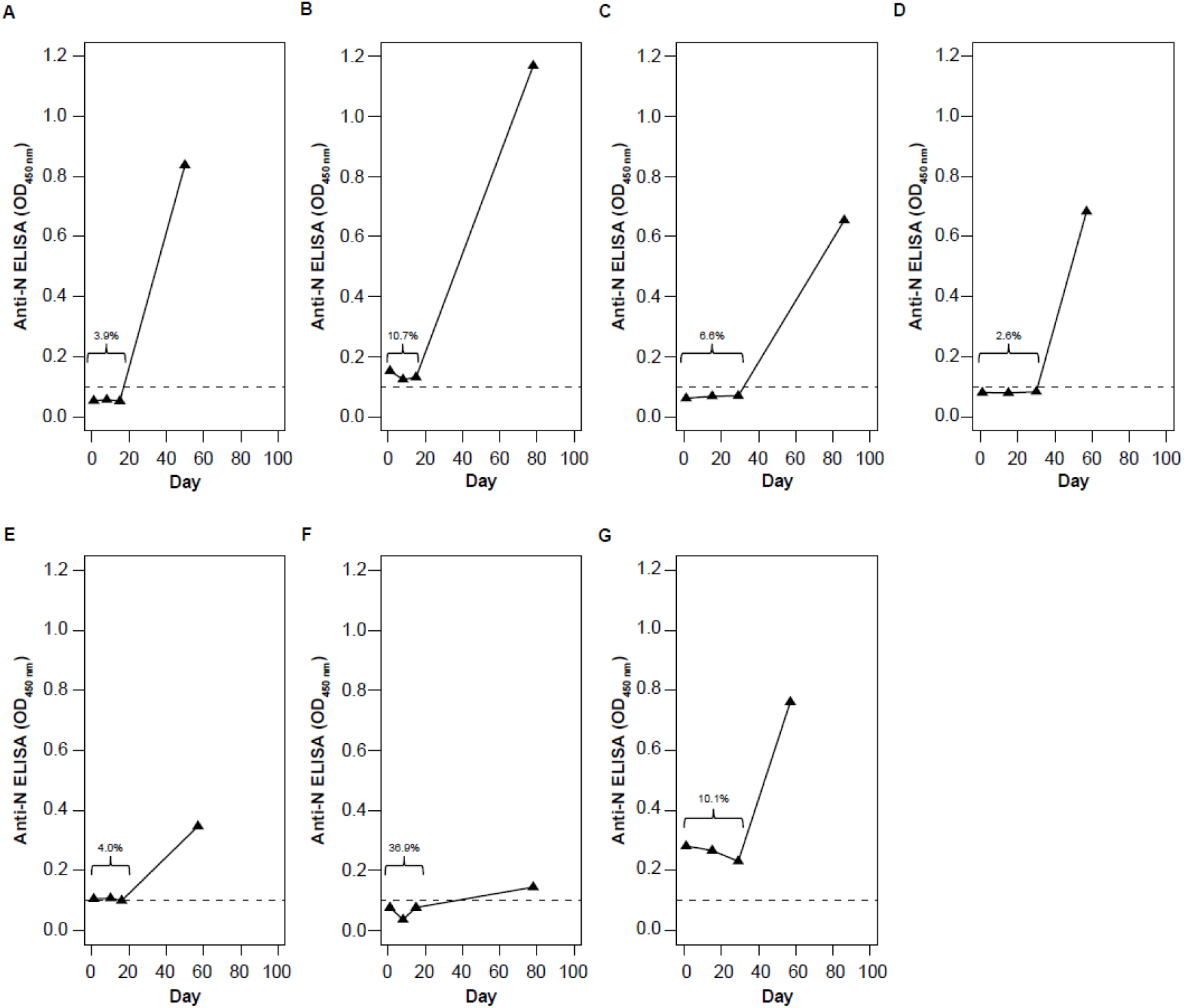
Stability of the anti-N signal in reference samples. Results of the anti-N ELISA for 7 individuals with three reference (pre-infection) samples. Day 0 represents the collection of the first reference sample; the interval (in days) between the collection of the first sample and the other samples is shown on the *x* axis. The first 3 dots on each panel show the results of reference samples and the last dot shows the result of the post-infection sample. The coefficient of variation of the results for the 3 reference samples is indicated on each graph.

### Testing the ratio-based approach in a real-world setting

The ratio-based approach was tested through a serosurvey that included 1618 plasma donors (mean age=54.0 years; females=32.8%) who donated during the Omicron wave, of which 1519 (93.9%) were included in the analysis conducted with the ratio-based approach. Using the ratio-based approach, the adjusted seroprevalence was 9.7% (95% CI=7.1%-12.4%) for the period that covered the beginning of the Omicron wave up until 01/17/2022-01/18/2022, 20.3% (95% CI=16.8%-23.8%) for that up until 02/14/2022-02/15/2022, and 27.4% (95% CI=23.8%-30.9%) up until 03/16/2022-03/18/2022. Using the conventional approach, the adjusted seroprevalence was 8.3% (95% CI=6.0%-10.5%) for samples collected between 01/17/2022 and 01/18/2022, 14.3% (95% CI=11.3%-17.4%) between 02/14/2022 and 02/15/2022, and 15.1% (95% CI=12.2%-18.0%) between 03/16/2022 and 03/18/2022 (**Figure 3**).

**Figure 3.**
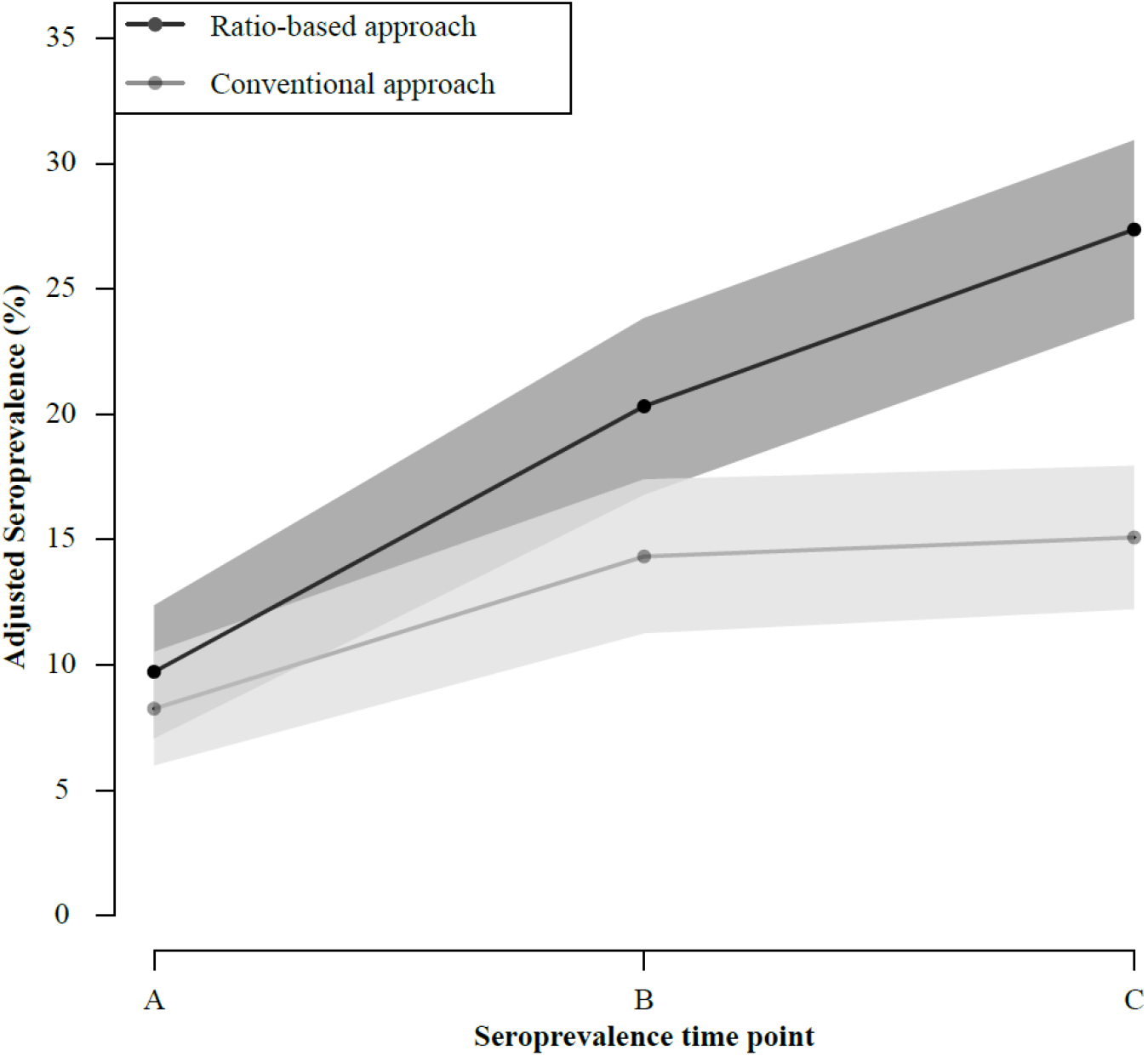
Adjusted seroprevalence obtained with the conventional and ratio-based approaches. The adjusted seroprevalence (age, sex, and regional distribution of the participants) was calculated using the ratio-based approach with samples covering 3 time intervals: A-C) since date of reference sample collection up to A) 17-18 January 2022, B) 14-15 February 2022 or C) 16-18 March 2022. The adjusted seroprevalence was also calculated using the conventional approach with samples collected at 3 different time points: A) 17-18 January 2022; B) 14-15 February 2022; C) 16-18 March 2022. The shaded areas around the lines represent the 95% confidence intervals of the seroprevalence estimates.

## DISCUSSION

This study presents a new approach (ratio-based approach) to assess the seroprevalence during a period of interest. The approach relies on longitudinally collected samples and determines seropositivity based on the difference between the result of a test sample and that of a reference sample collected earlier. In samples collected before and during the Omicron wave of the SARS-CoV-2 pandemic, the approach had a sensitivity of 95.2% among vaccinated individuals who had a breakthrough infection, which favorably compares with the sensitivity of 63.3% obtained with the conventional approach. Furthermore, at the latest time point considered in our analysis, this new approach estimated the adjusted anti-N seroprevalence at 27.4% during the Omicron wave, whereas this estimate was only 15.4% with the conventional single-sample approach. The conventional approach, as used in most (if not all) serosurveys, thus substantially underestimated the seroprevalence of anti-N antibodies in vaccinated individuals, even though it should theoretically reflect past infections since the onset of the pandemic, whereas the ratio-based approach measures seroprevalence only during a defined period of interest (the Omicron wave in the present study).

In addition to providing a high sensitivity, a cut-off of 1.5 for the anti-N ratio seemed adequate to reduce false positives that may arise due to the technical variability of the ELISA. The CV of negative controls (n=38 technical replicates) was 12.4%, and that of positive controls (n=38 technical replicates) was 11.3%, suggesting a cut-off lower than □1.3 (i.e., [100%+12%]/[100%-12%]) would be inadequate owing to the assay’s technical variability. The selected cut-off thus probably represents a good compromise between sensitivity and technical false positives.

The sensitivity of the conventional approach (63.3%) was higher than that reported in previous studies for vaccinated individuals (26.0%-40.4%).[15,16] Besides differences among assays, this discrepancy may be explained by the fact that the Omicron variant (for which vaccine efficacy is lower) was dominant during our study, whereas earlier variants (for which vaccine efficacy is higher) were dominant in previous studies.[19–22] As a result, individuals included in our study may have been exposed to higher viral loads than those included in previous studies, thereby increasing the likelihood of anti-N seroconversion.

The ratio-based approach estimated the adjusted anti-N seroprevalence at 27.4% during a period that covered the Omicron wave up until mid-March 2022. This figure is consistent with a recent modeling study, which estimated at 32% the proportion of Montreal residents who contracted SARS-CoV-2 between 12/01/2021 and 02/21/2022.[23] By contrast, the conventional approach estimated the adjusted anti-N seroprevalence at 15.1% in mid-March 2022. Together with the sensitivity data described above, these results demonstrate the extent to which the ratio-based approach captures a larger share of infections than the conventional approach.

This study has a few limitations. To begin, the true specificity of the ratio-based approach could not be assessed because of the unavailability of longitudinal samples from individuals ascertained not to have a history of SARS-CoV-2 infection (e.g., pre-pandemic longitudinal samples). However, the approach accounts for technical false positives by specifying a lower limit of absorbance below which test samples cannot be considered positive. In addition, the ratio-based approach requires access to at least 2 relatively recent samples per individual, which limits the pool of individuals that can be included in the analyses. Furthermore, the plasma donors who participated in PlasCov may not be representative of the general population. These individuals are generally healthier[24] and more of them are vaccinated against COVID-19 compared with the general population. Lastly, the legal age for plasma donation is 18 or older, and so children were not included in this study.

## CONCLUSION

This article describes a new and highly-sensitive approach (ratio-based approach) that successfully addresses the issue of vaccination in serosurveys. Relative to the conventional approach, the ratio-based approach captured a significantly greater proportion of vaccinated individuals with a recent history of SARS-CoV-2 infection (i.e., 95.2% vs. 63.3%). When tested in a real-world setting, this new approach also yielded a significantly higher anti-N seroprevalence than the conventional approach (i.e., 27.4% vs. 15.4%).

## Data Availability

All data produced in the present work are contained in the manuscript.

## ACKNOWLEDMENTS

The authors thank all the plasma donors who participate in the PlasCov biobank and the Héma-Québec personnel involved in sample collection and recovery from the biobank’s inventory.

