## Supplementary methods for "Development and use of a method based on the anti-N reactivity of longitudinal samples to better estimate SARS-CoV-2 seroprevalence in a vaccinated population"

### Supplementary Data

#### Supplementary Methods

*PlasCov Biobank*

The PlasCov biobank (https://www.hema-quebec.qc.ca/coronavirus/hema-quebec-en-contexte-de-pandemie/etude-plascov.en.html) is a biobank of samples obtained from regular apheresis plasma donations collected throughout Québec. PlasCov was established by Héma-Québec, the only blood operator in Québec, Canada. Sample collection started in April 2021 and is still ongoing. Informed consent was obtained at time of enrollment. The PlasCov biobank and the use of samples for the present study were approved by the Héma-Québec Research Ethics Board (REB # B6-2021-003 and # 2021-020). Participation rate of plasma donors in the PlasCov biobank is very high at 91%. The PlasCov biobank also includes information from the provincial database on vaccination and infection. PlasCov participants are adults (≥18 years) free of COVID-19 symptoms for ≥14 days before donation.

*Donor inclusion criteria*

To be included in the samples used to develop the ratio-based approach, PlasCov participants were required to meet the following criteria: (1) be aged ≥18 years; (2) be free of COVID-19-related symptoms within 14 days before their donation; (3) received ≥1 dose of a vaccine against SARS-CoV-2; (4) had PCR-confirmed SARS-CoV-2 infection during the Omicron wave, i.e., between 12/15/2021 and 03/20/2022; (5) made one donation before — but as close as possible to — 12/15/2021 (i.e., before the omicron wave; the "pre-Omicron donation"); (6) made another donation between 01/02/2022 and 04/20/2022 after the proven infection (the "Omicron donation").

For testing in a real-world setting, PlasCov participants were required to meet the following criteria: (1) be aged ≥18 years; (2) be free of COVID-19-related symptoms within 14 days before their donation; (3) made ≥1 donation in one of the following periods (all during the

Omicron wave): 01/17/2022-01/18/2022, 02/14/2022-02/15/2022, and 03/16/2022-03/18/2022.

*Conventional approach: seropositivity threshold, sensitivity, and specificity*

For the conventional approach, convalescent (n=52) and pre-pandemic plasma (n=66) collected in May 2020 were used to establish the optimal seropositivity threshold. An absorbance threshold of 0.350 maximized the sum of the assay's sensitivity (98.1%) and specificity (98.5%).
